# Urinary incontinence among pregnant women in Southern Brazil: a population-based cross-sectional survey

**DOI:** 10.1101/19011791

**Authors:** Yuan T. Hsu, Juraci A. Cesar

## Abstract

**Background:** Urinary incontinence (UI) is a frequent pathology that causes significant harm to the well-being and quality of life of pregnant women. This study aimed to measure the prevalence and to identify factors associated with the occurrence of UI during pregnancy in women living in the municipality of Rio Grande (RS), Southern Brazil, in 2016.

**Methods:** This is a cross-sectional population-based study that included all puerperae living in this municipality who had a child in one of the two local maternity hospitals between January 1 and December 31, 2016. The interviews were held at the hospital within 48 hours after delivery, when previously trained interviewers used a single, standardized questionnaire, seeking information on maternal demographic, behavioral and reproductive/obstetric history, as well as socioeconomic status of the household and care received during pregnancy and childbirth. The multivariate analysis followed a previously defined hierarchical model, using Poisson regression with robust variance adjustment and prevalence ratio (PR) as measure of effect.

**Results:** Among the 2,716 puerperae identified, 2,694 (99.2%) participated in this study. The prevalence of urinary incontinence in the gestational period was 14.7% (95%CI: 13.4%-16.1%). The lowest observed UI rate (8.3%) occurred among adolescent mothers (<20 years), while the highest occurred among those who reported frequent urinary urgency (39.2%). The probability of occurrence of UI, even after adjustment, was significantly higher among those who were older than 30 years old at current pregnancy, whose first pregnancy was before the age of 20 or after 30, who reached the end of gestation weighing 90 kg or more, who exercised regularly during pregnancy and who reported frequent urinary urgency during the gestational period.

**Conclusions:** Urinary incontinence showed a high prevalence in the study population. The identified risk factors can be well administered at primary health care level. The recommendation of regular physical exercise in pregnancy must be reviewed and better investigated with more robust designs because of possible facilitators for the occurrence of UI in this period.

## BACKGROUND

Urinary incontinence (UI) is defined as any involuntary loss of urine ^(1)^. It occurs in both genders and increases with age. Among women, its occurrence ranges from 40% to 60% and among men from 10% to 20% ^(2)^.

This higher occurrence among women is generally due to their reproductive life ^(3)^. Hormonal changes, enlargement of the uterus, pelvic floor changes during gestation and trauma suffered during delivery lead to involuntary loss of urine ^(4-6)^.

UI during gestation is an important predictor for its presence in subsequent pregnancies and at a later age ^(7)^. This makes it a chronic disease with a substantial deterioration of the quality of life, whether due to discomfort, the need for constant personal hygiene or insecurity, among others. At a later age, UI leads to isolation, which favors depression and more severe psychiatric conditions ^(8,9)^.

While very prevalent, UI has been poorly studied at the population level in Brazil. The few available studies are performed with a very small number of pregnant women, usually from a single health service, without any type of representativeness at the population level ^(10-12)^. In addition to preventing the establishment of actions and programs due to lack of knowledge of the real magnitude of the problem, this hinders prevention at the primary level of health care, which contributes to the persistence and severity of this disease, increases suffering and worsens the quality of life of these women, especially in the gestational period and in the older age. This study aims to measure the prevalence and to identify factors associated with the occurrence of UI in the gestational period among puerperae living in the municipality of Rio Grande (RS), Southern Brazil during 2016.

## METHODS

With a little more than 210 thousand inhabitants, the municipality of Rio Grande is located in the southern coastal strip of the state of Rio Grande do Sul (RS), about 300 km from the capital, Porto Alegre. Agribusiness, port activity and fishing are the main economic activities of the municipality. By 2015, its economy generated a gross domestic product (GDP) per capita of approximately US$ 12,000. In that same year, the infant mortality rate was 15 per thousand live births ^(13)^.

Data shown here derive from the 2016 Perinatal Study, which is part of a series of triennial cross-sectional surveys held in the municipality since 2007. These evaluations aim to monitor the quality of gestation and delivery assistance provided in this municipality.

To be included in this survey, pregnant women should reside in the municipality of Rio Grande (in urban or rural areas), must have had a child in one of the two local maternity hospitals (Santa Casa de Misericórdia or University Hospital) from January 1 to December 31, 2016 with a birth weight equal to or greater than 500 grams or with at least 20 weeks of gestational age.

The cross-sectional design was used, and mothers were approached only once in the maternity ward within 48 hours after delivery.

Regarding the sample size, two calculations were performed, namely, one to estimate prevalence and the other to identify associated factors. In the first case, using the expected prevalence of UI of 15%, error margin of 1.5 percentage points, level of significance of 95% and losses of 10%, the study should include at least 2,334 puerperae. Regarding the identification of associated factors, for alpha error of 0.05, beta error of 0.20, ratio between exposed and unexposed/exposed of 15/85, prevalence of disease among the unexposed of 9.8% and 1.6 risk ratio, at least 2,680 puerperae should be included in the study. These final values include 10% adjustment for eventual losses and 15% adjustment for control of potential confounders ^(14)^.

The outcome of this study was established by the event of urinary incontinence in the gestational period evaluated by the following question: “During this gestation, did you ever lose urine unintentionally?” A positive response would mean puerperae had UI during this gestational period.

The information about this study was collected through a single, pre-coded questionnaire applied by interviewers previously trained using tablets and the REDCap (Research Electronic Data Capture) application ^(15)^.

Three interviewers collected data. Two of them worked throughout the week and the other one on weekends and holidays. They visited these maternities of hospitals on a daily basis. When they arrived, they checked the number of births of the previous day in the register and visited all the rooms in the infirmaries allocated to puerperae. Once found, puerperae were asked to provide their place of residence. If in the municipality of Rio Grande, they would receive information on the study and, if they agreed to participate, they would sign two copies of the Informed Consent Form (ICF) and retain one copy. The other copy would be filed at the headquarters of this study. At least once a month, the team would meet with the project coordinator (JAC) to address any unclear issues, evaluate the application of questionnaires and review data collected via REDCap.

Questionnaires were uploaded daily through the REDCap Web platform ^(15)^ to the Federal University of Rio Grande (FURG) server at: www.redcap.furg.br. Then, data consistency was checked and immediately corrected. The consistency analysis, which included the categorization of variables and frequency verification, was performed using Stata statistical package version 12.0 ^(16)^.

Approximately 10% of the interviews were retaken in order to evaluate the quality of the data collected. This was done later by telephone or home visit. On that occasion, a summary questionnaire was applied. Kappa concordance index ranged from 0.68 to 0.89.

Results were expressed by the prevalence, and as a measure of effect, the prevalence ratio (PR), whose interpretation is similar to that provided to relative risk, in cohort studies, or odds ratio, in case-control studies. We also used a 95% confidence interval (95% CI) and p value of the trend test, and Wald test for heterogeneity ^(17)^. Crude and adjusted analysis was performed using Poisson regression, with robust adjustment for variance ^(18)^. The adjusted analysis was made based on a previously defined four-level hierarchical model ^(19)^. This adjusted analysis aims to eliminate the effect of confounding factors, that is, it separates the unique and exclusive effect of the variable in question on the endpoint, eliminating the effect of other variables that are not being tested.

These levels were used to determine the order of entry of the variables in the model. At the first level, demographic and socioeconomic variables (age, skin color, living with partner, schooling, household income and paid work during pregnancy) were included; the reproductive variable (age at the first pregnancy) was entered at the second level; variables related to prenatal and delivery care (number of prenatal consultations, trimester of onset of consultations, delivery type) and nutritional status (weight at the end of pregnancy) were added at the third level. The fourth and last level included variables related to habits and behavior (smoking, coffee consumption and regular physical activity in the gestational period) and morbidity (urinary urgency). The outcome was the event of urinary incontinence during pregnancy.

All the variables were taken to the multivariate model, and those with a value of p≤0.20 were maintained. Analyses were conducted in the Stata 12.0 program and the level of significance was 95%.

The research protocol was submitted and approved by the Health Research Ethics Committee (CEPAS) of the Santa Casa de Misericórdia of Rio Grande (file Nº 30/2015). In addition, data confidentiality, voluntary participation and the possibility of leaving the study at any time without need of justification were assured.

## RESULTS

The National Live Births Information System ^(20)^ and the Mortality Information System evidenced 2,716 births whose mothers lived in the municipality of Rio Grande. Of this total, 2,694 were interviewed, revealing a respondent rate of 99.2% (or a loss of 0.8%).

Table 1 shows the distribution of all puerperae by main characteristics studied. Practically, half of them were aged 25-29 years, two thirds were white, 83.6% lived with their partners, almost 70% had monthly household income of 1-3 minimum wages, about one quarter had completed at least elementary school, almost half (45.9%) engaged in paid work during pregnancy, 60.2% became pregnant during adolescence, 84.3% had at least six consultations in this gestation, 78.9% of them began consultations in the first trimester of pregnancy, 54.2% had a cesarean section, almost half of them (45.1%) weighed at least 80 kg at the end of gestation and 33.9% and 20.6% of them had coffee and/or smoked for at least one semester of pregnancy, respectively. About 6% had regular physical activity, 38.2% reported urinary urgency sometimes or frequently and 14.7% (95% CI: 13.4-16.1) reported having urinary incontinence.

**Table 1.** Prevalence of urinary incontinence according to some characteristics of puerperal residents in Rio Grande, Brazil, 2016.

| **Variables** | **Total (n)** |
| --- | --- |
| Maternal age (years) |  |
| 12–19 | 16.9% (456) |
| 20–29 | 49.7% (1,340) |
| ≥30 and over | 33.3% (898) |
| Maternal skin color |  |
| White | 67.0% (1,806) |
| Brown | 22.7% (610) |
| Black | 10.3% (278) |
| Lived with partner | 83.6% (2,252) |
| Household income in minimum wages |  |
| 0–0.9 | 8.5% (215) |
| 1–3.9 | 69.8% (1,775) |
| ≥4 | 21.7% (553) |
| Maternal schooling (full years) |  |
| 0–8 | 36.7% (990) |
| 9–11 | 39.8% (1,071) |
| ≥12 | 23.5% (633) |
| Engaged in paid work during pregnancy | 45.9% (1,237) |
| Age (years) at first pregnancy |  |
| 12–19 | 60.2% (924) |
| 20–29 | 35.2% (540) |
| ≥30 | 4.6% (71) |
| Number of consultations |  |
| 0–5 | 15.7% (422) |
| 6–11 | 72.5% (1,954) |
| ≥12 | 11.8% (318) |
| Started prenatal care in the first trimester | 78.9% (2,094) |
| Delivery type |  |
| Vaginal | 45.8% (1,234) |
| Cesarean | 54.2% (1,460) |
| Weight (kg) at the end of pregnancy |  |
| 40–69.9 | 28.7% (754) |
| 70–79.9 | 26.2% (688) |
| 80–89.9 | 22.1% (581) |
| ≥90 | 23.0% (607) |
| Drank coffee during pregnancy | 33.9% (912) |
| Smoked during pregnancy | 20.6% (341) |
| Engaged in regular exercise during pregnancy | 5.7% (154) |
| Had urinary urgency |  |
| Never | 61.8% (1,665) |
| Sometimes | 32.7% (881) |
| Often | 5.5% (148) |
| Prevalence of urinary incontinence | 14.7% (396) |
| Total | 100% (2,694) |

Of the incontinent women, 52.3% had stress incontinence, 18.4% urge incontinence and 29.3% mixed. Also, 8.8% of them started urinary loss in the first trimester of gestation, 27% in the second and 64.2% in the third trimester, and all of them had UI until the end of gestation.

Table 2 shows that the prevalence of UI ranged from 8.3% among adolescents (under 20 years) to 39.2% among those who reported frequent urinary urgency. In the adjusted analysis, the PR for puerperae aged 30 years or older was 2.05 (95% CI: 1.39-3.01) in relation to adolescents; mothers who had their first pregnancy aged 30 years or older, or before the age of 20, had PR=1.59 (95% CI: 1.01-2.51) and 1,36 (95% CI: 1.04-1.76), respectively, compared to those who had their first pregnancy at 20-29 years. In this same table, it is possible to verify that the greater the weight at the end of pregnancy, the higher the PR for UI occurrence. PR for the occurrence of UI among those weighing 90 kg or more was 1.63 (95% CI: 1.17-2.27) when compared to those who had a weight lower than 70 kg at the end of gestation. Finally, regular physical exercise during pregnancy and reporting frequent urinary urgency showed a PR of 2.49 (95% CI: 1.74-3.57) and 2.90 (95% CI: 2.10-4.00) in relation to those who did not exercise and did not report urinary urgency, respectively.

**Table 2.**
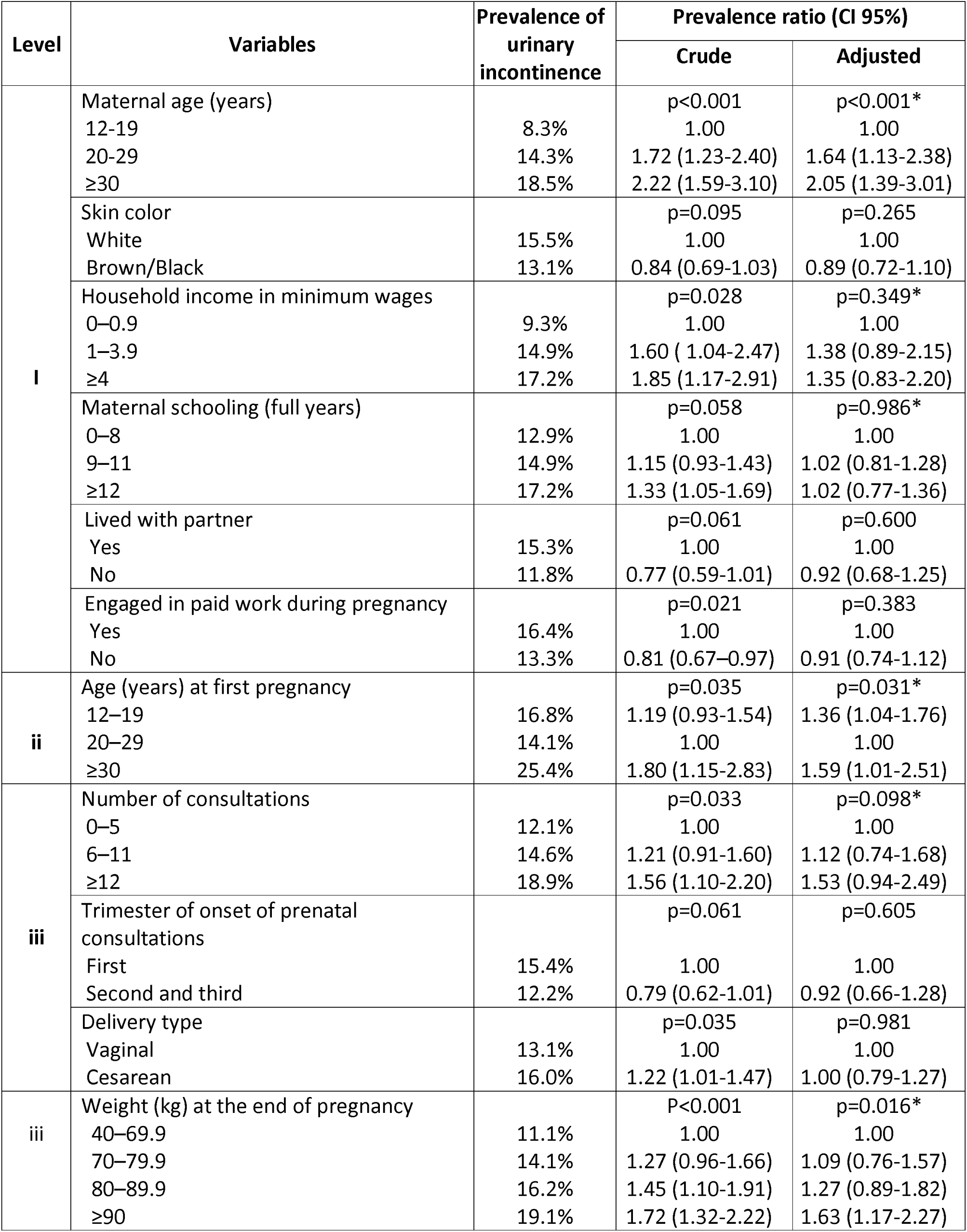
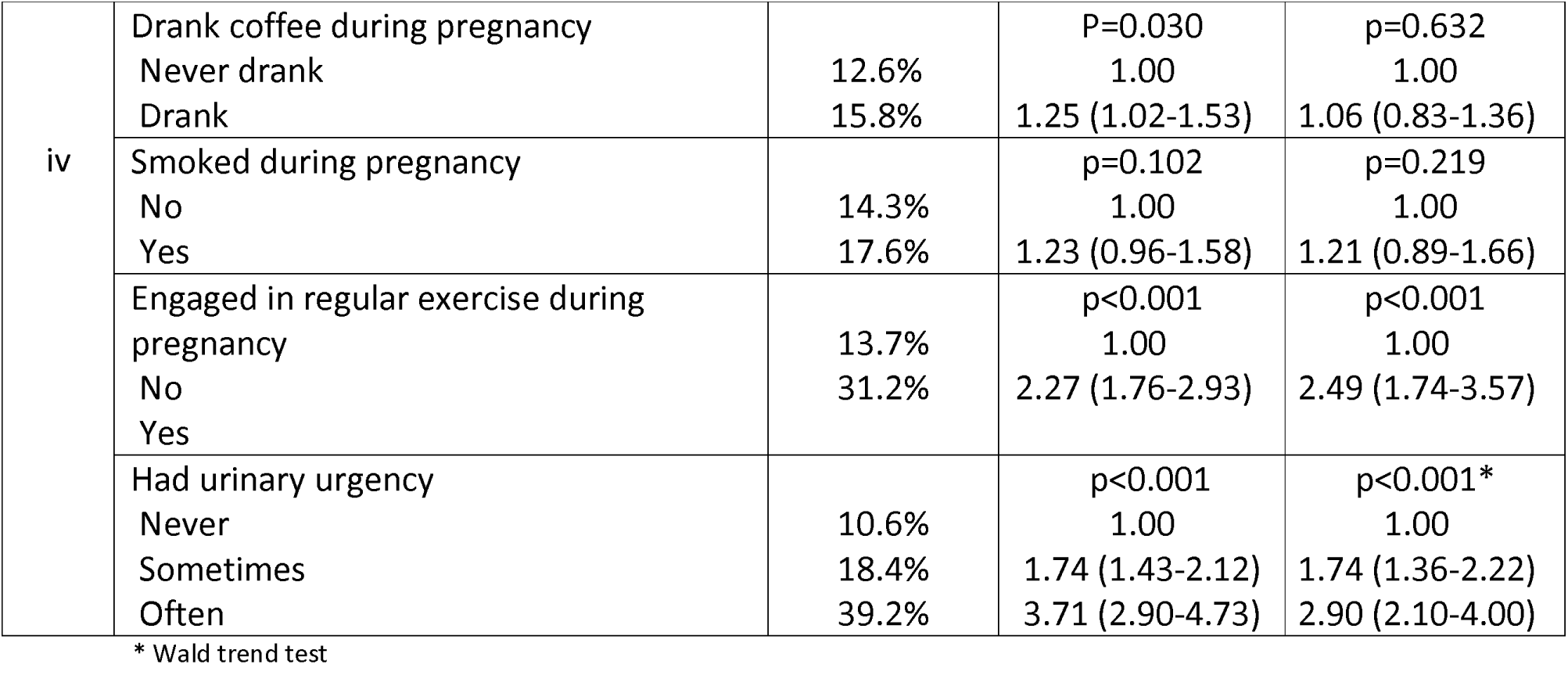
Prevalence of urinary incontinence by category and gross and adjusted analyzes according to hierarchical model. Rio Grande (RS), Brazil, 2016.

## DISCUSSION

This study found a prevalence of UI in the gestational period of 14.7%. It also showed that the probability of this disease, even after adjustment, is significantly higher among pregnant women who got pregnant and/or had a child in adolescence, who weighed 90 kg or more at the end of gestation, who performed regular physical exercises and who reported frequent urinary urgency during the gestational period.

The prevalence of UI found in this survey is low compared to other studies, ranging from 15% ^(21)^ to 71% ^(11)^. This enormous discrepancy arises from different characteristics of the participants, such as the inclusion of nulliparous alone, and the diagnostic criteria used, often based on a single question of the event of involuntary urine loss ^(22-24)^.

Maternal age is an inexorable marker of the occurrence of UI. The greater the age, the greater its prevalence. This is due to the loss of innervation and gradual reduction in the contraction capacity of muscle fibers and increased permeability of the urethral sphincter ^(25)^. This leads to a lower pressure of its closure ^(26)^, resulting in the involuntary loss of urine. A recent systematic review conducted in the European population found OR=1.4 (95% CI: 1.3-1.5) for UI among those 35 years of age or older compared to younger age ^(21)^. A similar result was found in this study. Mothers aged 20-29 years and 30 years or older showed a PR for UI of 1.64 (95% CI: 1.13-2.38) and 2.05 (95% CI: 1, 39-3.01), respectively. This evidences the strength of the variable age as a risk factor for this condition.

Maternal age at the time of the first gestation was also significantly associated with the probability of UI in the studied population. A similar finding was found in a cross-sectional study conducted in Norway with about 11,000 women ^(27)^. In this study, women with gestation before 25 years of age showed a prevalence of UI of 23% versus 28% among those who had a child at a later age (p> 0.001). In this study from Rio Grande, having a child at 20-29 years showed the lowest risk of UI compared to those who had it before the age of 20 or after the age of 30. This is probably due to pelvic floor trauma at younger ages and loss of muscle fibers and urethral sphincter pressure at later ages.

In this study, weighing over 90 kg at the end of gestation showed PR=1.63 (95% CI: 1.17-2.27) in relation to the others. The relationship between body mass index (BMI) and UI is usually directly proportional. This risk factor is already well established ^(28,29)^. This may not only be due to the relationship between weight and height, but also that the gestational period shows an increased bone density and peripheral edema, in addition to the influence of hormonal factors and fetal weight. This set of factors may be responsible for increased weight gain being an important risk factor for UI.

In Rio Grande, regular physical exercise, even after adjustment, appeared as a predisposing factor to UI in relation to the other pregnant women. This is even more worrying because, during pregnancy, it is common for obstetricians to indicate physical exercise, called “fitness”, for pregnant women. They claim the benefits of this practice to maternal-fetal health and, because of this, are included in several guidelines as a healthy measure for gestation ^(30)^.

It is well known that, even at a young age, elite female athletes have a higher prevalence of UI. This prevalence can affect about half of them^(31)^. A Norwegian study conducted among academy instructors, including Pilates and Yoga teachers, found a prevalence of UI of 26.4% among instructors with a mean age of 32.8 years (± 8.3). This rate is very similar to that observed in the general female population ^(32)^. These data suggest that physical exercises can overload the pelvic floor, thus increasing the likelihood of UI. In the case of pregnant women, who already have an overload, this is even more serious. Hence the need for this indication to be very well defined, mentioning exactly which exercises, how often at what time of gestation they can be performed. Otherwise, this indication may favor UI. It should be noted, however, that when the exercise is directed to the pelvic floor musculature training (PFMT), it has been effective in reducing the occurrence of UI, with RR=0.71 (95% CI: 0, 54-0.95) compared to those who did not perform this type of training ^(33)^. Also on this subject, a meta-analysis showed that PFMT, by reducing labor time, especially in the first and second stages reduces pelvic floor trauma and, therefore, can prevent the occurrence of UI ^(34)^.

About 40% of pregnant women in this study reported urinary urgency, that is, a sudden sensation that makes it very hard to postpone urination. Of these, about six percent referred to this condition as very frequently. The PR for the probability of UI of this group among those who did not report urinary urgency was very high at 2.90 (95% CI: 2.10-4.00). The association between urgency and incontinence is so close that it was set in the immediate upper level of the endpoint, showing the relevance of this condition to the occurrence of UI.

Urinary urgency is a very frequent problem among pregnant women. A Brazilian study found prevalence of this condition in 44% of the participants^(35)^. The main reason for urinary urgency is the increased blood volume and the effect of circulating hormones during pregnancy ^(36)^, but this needs to be better clarified.

When interpreting these results, it is necessary to consider that this is a cross-sectional study. Therefore, caution should be exercised when interpreting some associations because the exposure and outcome variables were collected at the same time. However, we could not find in the literature a more robust design that has a work with such an expressive number of pregnant women as this one.

## CONCLUSIONS

This population-based study showed that urinary incontinence is a common disease among pregnant women. In addition, it confirmed the findings of other investigations for maternal age and urinary urgency as a risk factor for this ailment and suggests that the variables “age at first gestation”, “weight over 90 kg at the end of pregnancy” and “engaging in regular physical exercise in this period” may be causally associated with this disease as well. As a result, it is recommended that professionals providing prenatal care pay attention to these factors, as well as we suggest that, when using a more robust design such as a cohort, these three variables be included in future research on this topic.

## Data Availability

I declare that the data is available

## LIST OF ABBREVIATIONS

BMI: Body Mass Index
CEPAS: Health Research Ethics Committee
GDP: Gross Domestic Product
ICF: Informed Consent Form
OR: Odds Ratio
PFMT: Pelvic Floor Musculature Training
PR: Prevalence Ratio
REDCap: Research Electronic Data Capture
RS: Rio Grande do Sul - Name of the State
UI: Urinary Incontinence

## DECLARATIONS

### Consent for publication

not applicable

### Availability of data and materials

the datasets used and analysed during the current study are available from the corresponding author on reasonable request.

### Competing interests

I confirm that I have read BioMed Central’s guidance on competing interests and none of the authors have any competing interests.

### Funding

About 80% of this amount was paid by the students involved in this study and by their supervisor. The rest was paid by CNPq (Conselho Nacional de Pesquisa - Ministry of Science, Technology and Innovation) via the productivity grant of one of the participants. The FURG University Hospital provided a meal to all interviewers throughout the data collection period, while the FURG Graph provided all free and informed consent terms.

### Authors’ contributions

Hsu Yuan Ting – the main author. Juraci Almeida Cesar – doctoral supervisor.

## Acknowledgements

not applicable

## Notes

### Competing Interest Statement

The authors have declared no competing interest.

### Funding Statement

About 80% of this amount was paid by the students involved in this study and by its coordinator. The remainder was paid by CNPq through a productivity grant from one of the participants. The University Hospital provided meal to all interviewers throughout the data collection period, while University Printing provided all informed consent (ICF) terms.

